# Comparative analytical sensitivity evaluation of five mpox point-of-care assays for mpox clades Ia, Ib, IIa and IIb

**DOI:** 10.64898/2026.01.17.26344231

**Authors:** Kenneth Adea, Camille Escadafal, Devy M. Emperador, Emmanuel Agogo, Megan O’Driscoll, Placide Mbala-Kingebeni, Laurent Kaiser, Meriem Bekliz, Isabella Eckerle

## Abstract

**Backgrounds:** Following an increase in mpox clade Ib cases in several African countries, the World Health Organization recognized mpox as a public health emergency of international concern, highlighting the need for reliable and accessible diagnostic tools. As several mpox clades are co-circulating in endemic countries, the analytical sensitivity of diagnostic assays should be evaluated for all of them in a comparative manner.

**Methods:** This study evaluated the analytical sensitivity of three rapid tests (Flowflex Monkeypox virus rapid test from ACON Biotech, Monkeypox antigen rapid test from Assure Tech, and Standard Q Monkeypox Ag Test from SD Biosensor) and two point-of care molecular tests (SD Biosensor’s M10 and Cepheid’s Xpert MPOX assays) using serial dilutions of viral stocks from the four clades of Monkeypox virus (MPXV): Ia, Ib, IIa, and IIb.

**Findings:** Upon our analytical comparative benchmarking, the three rapid tests demonstrated comparable analytical sensitivity for all four clades, with a limit of detection of approximately 1,000,000 DNA copies/mL or 1,000 FFU/mL. The two molecular tests demonstrated also comparable analytical sensitivity to an in-house PCR assay for all four clades, detecting concentrations down to 10-100 DNA copies per mL, corresponding to a non-detectable titer of infectious particles. The Xpert assay detected the Clade Ib strain only through its orthopoxvirus target and not its MPXV target, and none of the assays could distinguish between MPXV clades.

**Interpretation:** No differences in sensitivity was observed across MPXV clades neither for Ag-RDTs nor for molecular POCTs. However, the potential of simple, affordable tests, such as Ag-RDTs, is limited by poor sensitivity while the use of highly sensitive POC molecular platforms remains limited by their cost. To date, the lack of accurate, affordable POC MPXV-specific assays with potential to differentiate clades, limits diagnostic capacities as well as our understanding of the virus and its epidemiology.

**Funding:** This work was supported by FIND and internal funds of the Centre for Emerging Viral Diseases. Mpox diagnostic tests were provided by FIND and the World Health Organization (WHO). MOD was supported by Schmidt Science Fellows, in partnership with the Rhodes Trust.

## Introduction

Monkeypox virus (MPXV) is an enveloped double-stranded DNA virus belonging to the family *Poxviridae* in the genus *Orthopoxvirus* (1). It is the causative agent of mpox disease, which manifests with symptoms similar to those of mild cases of smallpox. The first documented human case of the disease occurred in 1970 in the Democratic Republic of Congo (DRC)(2). Following the multicountry outbreak of mpox in 2022, the World Health Organization (WHO) formally declared a Public Health Emergency of International Concern (PHEIC)(3). Subsequently, more than 91’000 confirmed cases were reported in 116 non-endemic countries, with notable occurrences in the United States, Brazil, and Spain (4). MPVX is classified into two clades: clade I (formerly the Congo Basin strain) and clade II (formerly the West African strain). Clade II is further divided into two subclades, clade IIa and clade IIb, with clade IIb accounting for most of the circulating strains outside Africa. The MPXV clades I and IIa are endemic to central and western Africa, respectively. Since the beginning of 2023, the number of mpox cases worldwide had decreased, but a new outbreak began in late 2023 in the DRC caused by a new variant of MPXV clade I, named clade Ib, to distinguish it from previous clade I variants, now identified as clade Ia (5, 6). The increase in cases in the DRC, and then in a growing number of African countries, led to this new outbreak being recognized as a public health emergency of continental security by the Africa Centers for Disease Control and Prevention and renewed as a PHEIC by the WHO in August 2024 (7, 8).

The gold standard method for the diagnosis of MPXV infection is polymerase chain reaction (PCR) based on the detection of MPXV DNA in muco-cutaneous, oropharyngeal, or blood specimens (9). Other molecular approaches based on isothermal DNA amplification, as well as serological methods based on immunoglobulin (IgG or IgM) or antigen detection, have been developed and documented in the literature but have demonstrated lower performance compared with PCR (10, 11, 12). Given the poor and scarce data on the performance of antigen tests, as of August 2025, there is no Emergency Use Listing (EUL) process for MPXV antigen rapid diagnostic tests (Ag-RDTs), while there is one for molecular tests (13). However, most current PCR methods require specialized personnel, expensive equipment and consumables, and long analysis times (14, 15). Consequently, there is a critical need for affordable and easy-to-use diagnostic tests for mpox diagnosis, which would allow for wider access and decentralization of testing, notably in low-income countries or underserved communities (16).

According to the latest FIND test directory data, there are 12 “*research use only*”, and 10 regulatory approved point-of-care (POC) molecular assays for MPXV diagnosis (17). Most of these commercially available tests are cartridge-based assays using near-POC platforms rather than true-POC (portable, battery-powered). Information on the potential impact of the different MPXV clades on test performance is limited, and experimental data on the analytical sensitivity of POC molecular tests for clade Ib are not available.

In 2022, FIND, a global nonprofit that connects partners to spur diagnostic innovation and access, launched an initial open call to mpox test manufacturers and selected five tests for evaluation (18). Although some clinical and analytical performance data on the three Ag-RDTs have been recently published, the analytical sensitivity testing only included clade IIb viral dilutions (19, 20). There is a need for comparative analytical sensitivity data on the POC molecular tests that represent all circulating MPXV clades, including the newly emergend Ib that was not yet recognized when the majority of commercial diagnostic tests for Mpox became available (21). In this study, we evaluated the analytical sensitivity of two near-POC molecular assays and three Ag-RDTs using viral dilutions of cultured isolates for each of the four MPXV clades: Ia, Ib, IIa, and IIb.

## Results

### Analytical sensitivity of POC molecular assays for all four MPXV clades

The analytical sensitivity of two near-POC molecular assays was evaluated for all four clades: the Standard M10 assay from SD Biosensor and the GeneXpert assay from Cepheid.

The individual triplicate test results for each molecular assay across a range of viral dilutions are reported in Tables S1, S2, S3, and S4, for Clades Ia, Ib, IIa, and IIb, respectively. Figure 1 shows the overall results of both assays and all four clades, representing the last dilution that was detectable by all three replicates, with the unit set as DNA copies/mL as quantified by a diagnostic orthopox PCR assay (22).

**Figure 1.**
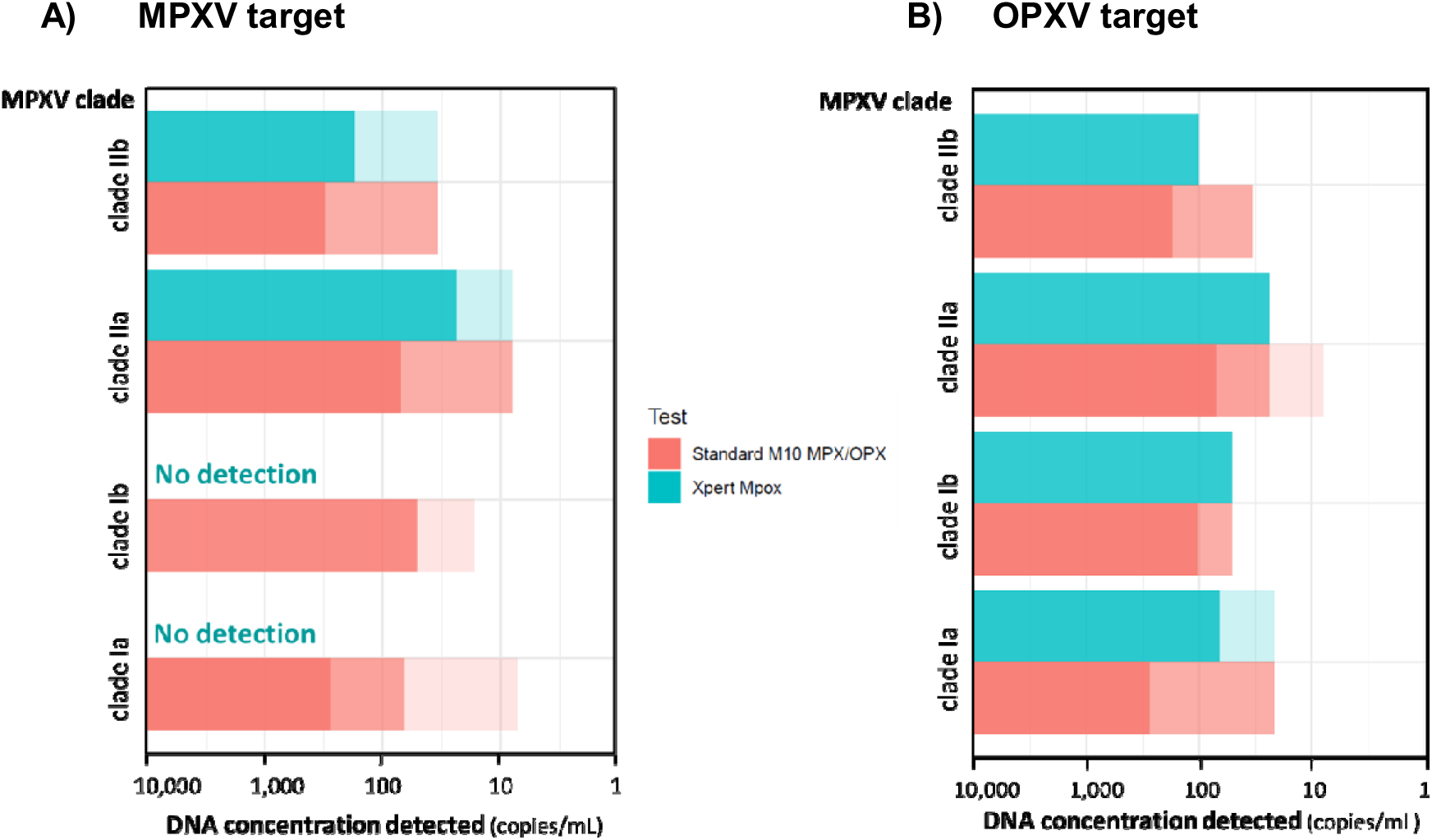
Analytical sensitivity of the two POC molecular tests to detect viral dilutions of MPXV clades Ia, Ib, IIa, and IIb. The bars represent the viral concentrations in DNA copies per mL. The darker shade bars represent the concentrations detected by all three replicates, the medium shade bars represent the concentrations detected by only two replicates, and the lighter shade bars represent the concentrations detected by only one replicate. A) Results of the MPXV target assays; B) Results of the OPXV target assays.

No significant differences were observed in the analytical sensitivity between the two molecular assays or the four different clades. The last detected concentrations ranged from approximately 10 to 100 DNA copies/mL for both tests and all clades. Based on the focus forming assay (FFA) titration results that we obtained, concentrations in this range did not allow culture or detection of infectious viral particles (Figure S1).

Importantly, clade Ia and Ib dilutions were only detectable by the OPXV target and not by the MPXV target of the Xpert assay, but were detectable by the MPXV target of the SD Biosensor assay.

The error rate (i.e., the percentage of tests for which the output was an error message instead of a valid result) for the Xpert and M10 assays was 5.4% and 0%, respectively. The intra-assay repeatability was assessed using triplicate tests for each sample. Discordant results were observed in 22 of the 96 samples tested (22.9%). These results were mostly observed when the concentrations were low, but not exclusively.

### Analytical sensitivity of Ag-RDTs for all four MPXV clades

The analytical sensitivity of Ag-RDTs for detecting cultured MPXV clade Ia, Ib, IIa, and IIb viruses was evaluated. Three Ag-RDTs were evaluated: (i) Flowflex Monkeypox Virus Rapid Test from ACON Biotech (hereafter referred to as Flowflex), (ii) Monkeypox antigen rapid test from Assure Tech (Assure Tech), and (iii) Standard Q Ag Test from SD Biosensor (Standard Q) (Table 1).

**Table 1.**
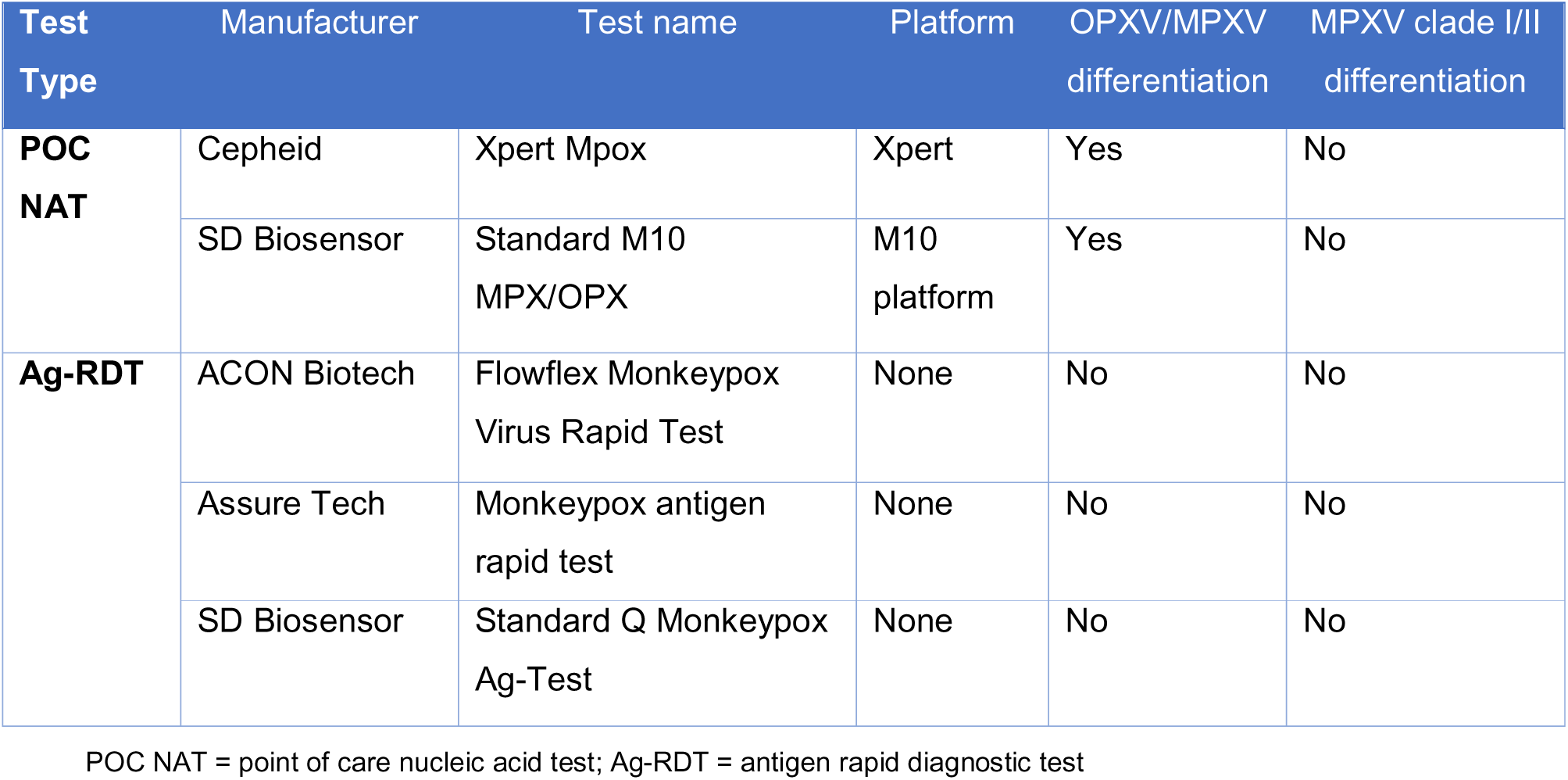
Overview of the molecular POC and Ag-RDT assays evaluated in this study.

Individual test results for each Ag-RDT and each triplicate are reported in Tables S5, S6, S7, and S8, for Clades Ia, Ib, IIa, and IIb, respectively. Figure 2 presents the overall results of the three Ag-RDTs and all four clades as the last dilution detected by all three replicates. The unit is set as DNA copies applied to the Ag-RDT to account for the different input volumes applied to each test: 100µL of buffer/dilution mixture for Standard Q and 200µL for Assure Tech and FlowFlex.

**Figure 2.**
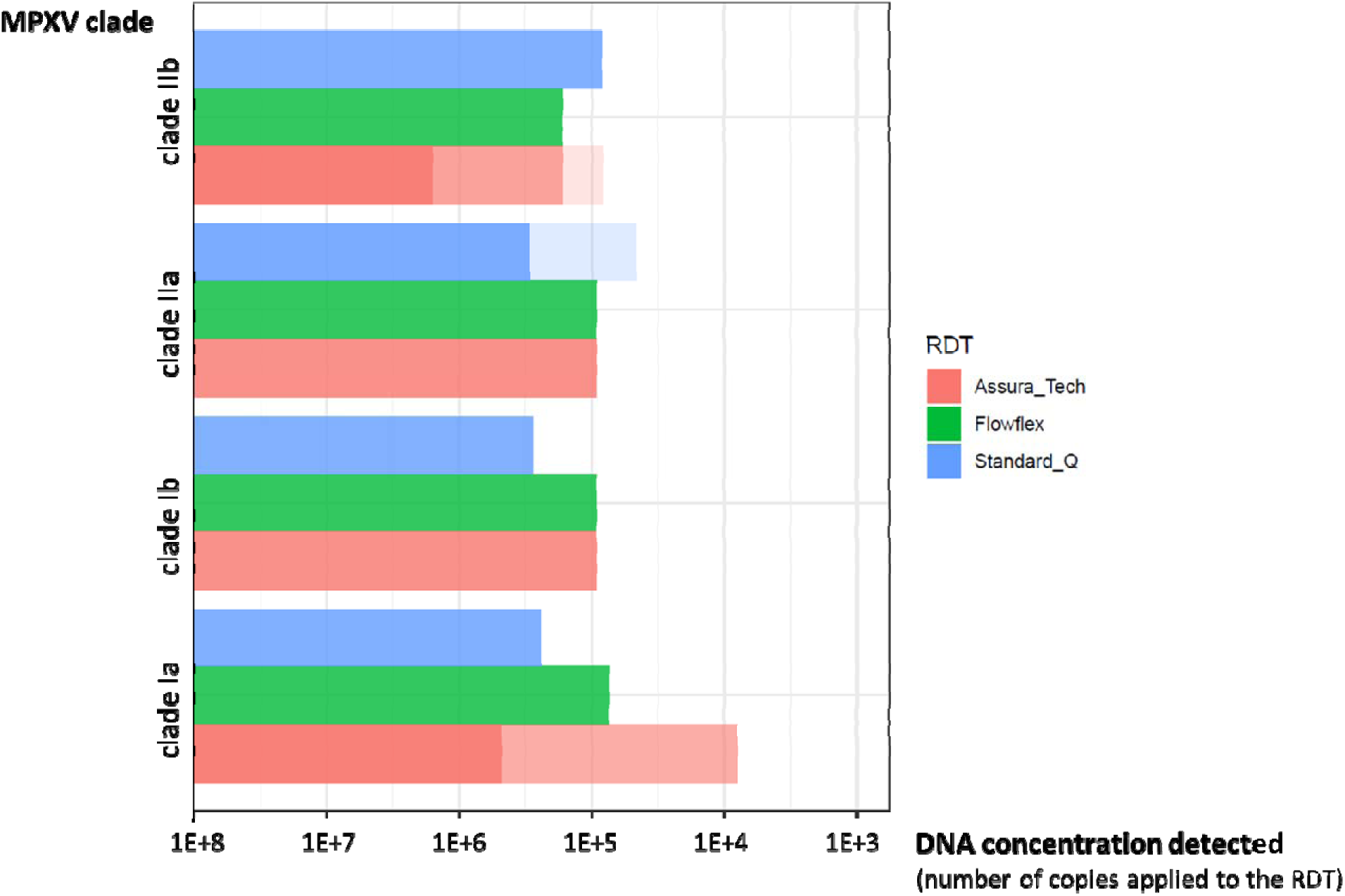
Analytical sensitivity of the three Ag-RDTs in detecting viral dilutions of MPXV clades Ia, Ib, IIa, and IIb. The bars represent the viral concentrations detected in DNA copies applied per test. The darker shade bars represent the concentrations detected by all three replicates, the medium shade bars represent the concentrations detected by only two replicates, and the lighter shade bars represent the concentrations detected by only one replicate.

The analytical sensitivity of Ag-RDTs was significantly lower than that of the molecular tests. This low performance was observed across the three Ag-RDTs and four MPXV clades. None of the Ag-RDTs could detect concentrations lower than 10^5^ DNA copies/ml, which is equivalent to fewer than 10^4^ DNA copies per test. Viral titration results of each clade showed that infectious viral particles are present in dilutions with a DNA copy concentration down to 10^3^ copies/mL (Figure S1).

The rate of invalid tests (i.e., tests with no visible control line) was 0% for all three Ag-RDTs. The intra-assay repeatability was assessed using triplicate tests for each sample. Discordant results were observed in seven out of 108 samples (6.5%), only when the concentrations were low and the test bands were faint. No discordance was observed between the results of the two independent readers for each Ag-RDT.

## Discussion

This study generated benchmarking data on the analytical sensitivity of two POC molecular and three antigen rapid diagnostic assays for the detection of the 4 different MPXV clades, providing insight into the potential impact of different MPXV clades on the performance of POC assays.

There were no significant differences in performance between the two POC molecular tests or between each of the three Ag-RDTs and the results revealed no significant differences in analytical sensitivity among the four MPXV clades, regardless of the assay type. However, the Xpert MPX/OPX assay produced false-negative results for the MPXV target for both clade I strains. This assay is used in countries currently facing clade I outbreaks, where MPX-negative/OPX-positive results are interpreted as MPXV clade I (23). However, this assay cannot be considered a clade-specific assay and can lead to misinterpretation of the results.

Furthermore, the results showed a major difference in analytical sensitivity between the POC molecular tests and Ag-RDTs, with the molecular tests detecting down to 10^1^-10^2^ DNA copies/mL, whereas the Ag-RDTs could not detect DNA concentrations below 10^5^ copies/mL under our study conditions.

Several studies have indicated high viral loads in lesion swabs, which is the recommended sample type for mpox diagnosis. In a systematic review of nine articles, the pooled mean Ct value of skin samples positive for mpox is 21.71 (95% CI: 20.68–22.75)(24). Oropharyngeal swabs (OPS) are also used for diagnostic purposes and, viral loads are reported to be significantly lower than those in skin lesions (Ct = 27.4, 95% CI: 26.1-31 for OPS; Ct = 21.2, 95% CI: 20.5–22.3 for skin lesions)(25) but still below a Ct of 30. Two studies correlating viral load and infectious virus titers in clinical specimens suggest that specimens with a Ct value greater than 35 (estimated to be equivalent to ≤ 4,300 DNA copies/mL) have no or marginal infectivity (26, 27). These clinical data suggest that the two POC molecular assays we evaluated would detect all infectious samples (i.e., all samples with over 4,300 DNA copies/mL) as mpox positive, whereas the three Ag-RDTs would likely miss a significant proportion of infectious samples. However, the two types of tests have different use cases, and WHO published two different product target profiles for mpox diagnostic tests, one for molecular tests for use in laboratory settings, and one for lateral flow rapid tests for use in community settings. The use of rapid tests with a lower sensitivity may still be relevant as a rule-in test in high prevalence settings considering their quick turnaround time and higher accessibility. Modelling studies in other contexts, e.g. the use of Ag-RDTs for Ebola virus disease have shown that even the use of rapid tests with suboptimal sensitivity can be beneficial in an outbreak by limiting time to isolation when used as a rule-in test (28).

Although increased sensitivity is generally considered an important quality in diagnostic tests, the detection of low nucleic acid concentration may not always be clinically significant. Multiple studies have reported the detection of MPXV DNA in air and surface samples from mpox patient care units (29, 30). Although these findings reinforce the importance of rigorous infection control, positive test results from sensitive molecular assays may not always be clinically relevant. Test results must be considered alongside clinical information to prevent patients from being falsely identified as cases of mpox, which could lead to them receiving inappropriate care as well as inaccurate surveillance data being recorded.

In terms of test usability, the error rate or invalid test rate (i.e., the percentage of tests for which the output was an error message or Ag-RDTs with no visible control line) was 0% for the three Ag-RDTs and the M10 assay, whereas the Xpert assay had an error rate of 5.4%. Although the naked-eye reading of Ag-RDTs provides a more subjective interpretation than automated platforms, the inter-assay repeatability was higher for Ag-RDTs (93.5% vs. 77.1% of concordant results) than for POC molecular assays. All three Ag-RDTs were easy to use, and the results were available 15 minutes after the sample was added to the cassette. The molecular POC assays are also user-friendly. The user simply adds the sample to a cartridge and loads it into the system. Both assays have a turnaround time of approximately 1 hour. Neither platform is a true-POC system, but rather a near-POC system, as a three- to four-module system weighs more than 10 kg. The GeneXpert platform is slightly less bulky than the M10 platform but requires a computer connection. These near-POC assays are not compatible with field testing in resource limited settings without laboratory infrastructure.

Our study has a few limitations. As we aimed to provide a benchmark for analytical sensitivity comparing the all MPXV clades, we did not study the analytical specificity of the assays. It is known that orthopoxviruses share genome homology and therefore there is a theoretical risk of cross-detection of other orthopoxviruses by molecular or even antigen-based methods. However, analytical specificity performance data has been provided by all the manufacturers of the tests we evaluated and these have shown no cross-reactivity. Moreover, it was reported by previous mpox test performance studies that sensitivity is the primary limiting factor for test performance and not specificity (20,31,32). Lastly, to date, no other Orthopoxviruses were reported co-circluating in the DRC.

Furthermore, our reference test is based on the PCR protocol published by Scaramozzino et al. (14). This method proved to be less sensitive than the tests we are evaluating, resulting in an estimated value of the last DNA concentrations detected by the evaluated tests. The reference test was designed to detect all orthopoxviruses and was not optimized for the different MPXV clades. The sequences of our four MPXV viral stocks showed one mismatch in the reverse primer region and one additional mismatch in the probe binding region for clade Ia and Ib viruses. Therefore, the assay may exhibit slightly lower sensitivity for clade I vs clade II, even though the probe is long (33 bp) and the mismatch is toward the end of the probe, which could lead to an underestimation of DNA concentrations for clade I viral dilutions.

Another limitation was observed when MPXV serial dilutions were prepared. We aimed for a dilution factor of three between each dilution step. Despite careful vortexing and pipetting up and down at each dilution step, achieving consistent dilution factors was difficult, particularly at lower viral concentrations. This issue may be due to the aggregation of multiple viral particles and could be prevented by using a different dilution buffer to adjust parameters such as pH or ionic strength. Although viral transport medium (VTM) is commonly used to transport and store swab samples for mpox PCR testing, VTM has been reported to have inhibitory effects on the detection of antigens (12). Therefore, PBS was used in our study as a dilution buffer across all validations and not VTM in order not to add an additional bias but only study the analytical sensitivity to the virus clade.

Most importantly, our evaluation does not replace clinical performance testing in a real-world setting using characterized clinical samples. Due to ethical, logistical and regulatory restrictions, we did not have access to a panel of clinical samples from all four MPXV clades to include them to our validation. Considering that the circulation of the clades occurs in different geographic regions and that the zoonotically transmitted clade Ia and IIa viruses are rare, no such comparative study across all four clades would be possible in a real-world setting. Two published performance studies have evaluated separately the 3 Ag-RDTs and the 2 POC molecular plaftorms included in our study, but these present analytical and clinical data from one clade only (20,31). It remains important to underline that more clinical studies are needed to complement analytical results such as ours and provide a comprehensive understanding of the performance of these tests in the settings where they are needed the most.

Overall, this study provides valuable insight into the analytical sensitivity of mpox POC assays and potential clade-specific variations that may affect the accuracy of diagnostic tests, thereby affecting patient care and outbreak management. This knowledge is critical for public health professionals when implementing diagnostic strategies, especially in regions where mpox is endemic and more than one clade may be circulating.

The Xpert MPX/OPX assay could not detect clade I samples through its MPX target, and none of the evaluated assays could distinguish between MPXV clades. This highlights the importance of developing MPXV-specific assays capable of distinguishing between clades and improving our understanding of the virus and its epidemiology, as sequencing and clade-determination is only possibe for a small subset of samples in such remote settings.

Rapid, affordable, and sensitive mpox diagnostic tests are essential for controlling global outbreaks. However, the wider potential of molecular platforms for decentralized testing is currently limited by their cost. Conversely, the potential of simple, affordable tests, such as Ag-RDTs, is limited by their poor sensitivity but might still be useful in the context of high prevalence and lack of other diagnostic tools.

## Materials & Methods

### Index tests

The five index tests included in this study are listed in Table 1. FIND selected two POC molecular assays and three Ag-RDTs based on criteria including ability to detect MPXV clade I and II strains, available analytical and clinical performance data and company strength (19). The GeneXpert assay was listed under the WHO EUL procedure in October 2024 (33).

### Reference test

The reference test selected for this study was the PCR described by Scaramozzino-N *et al*. targeting the open reading frame (ORF) A30L of orthopoxviruses (14). Viral DNA was extracted using the NucliSens easyMAG kit (BioMerieux), and real-time PCR was performed using the *Taq* DNA Polymerase kit (Invitrogen) in a CFX96 Thermal Cycler (BIORAD).

### Virus and cell culture

Vero-E6 cells (ATCC CRL-1586) and Vero-E6-TMPRSS (engineered Vero-E6 overexpressing TMPRSS2 protease, provided by National Institute for Biological Standards and Controls, NIBSC, Cat. Nr. 100978) were cultured in DMEM GlutaMAX I medium supplemented with 10% FBS, 1x non-essential amino acids, and 1% penicillin-streptomycin (all reagents from Gibco, USA).

The Spiez Laboratory in Switzerland shared the MPXV clade Ia, IIa, and Ib isolates used in this study. Clade Ia and Ib isolates were shared via the WHO BioHub System (reference numbers 2023-WHO-LS-008 and 2024-WHO-LS-003, respectively), which provides a reliable and transparent mechanism for voluntarily sharing novel biological materials. Clade IIb was obtained from residual clinical samples collected from a patient presenting at the University Hospitals of Geneva (HUG), under the HUG’s general informed consent that allows the use of anonymized leftover materials for virus isolation. All four viral isolates were fully sequenced, and references to the sequences are available in Table S9.

The MPXV clades Ib, IIa, and IIb strains were isolated and propagated on Vero-E6 cells, and clade Ia on Vero-E6 TMPRSS2 cells. For virus stock preparation, only the supernatant was harvested. All four clades were titrated on Vero-E6 cells. All experiments involving live, infectious MPVX were approved and adhered to the standard operating procedures of our Biosafety Level 3 (BSL-3) facility.

### Viral load quantification by qPCR

After DNA extraction, viral DNA was quantified using quantitative real-time PCR (qPCR) with a *Taq* DNA Polymerase kit (Invitrogen) in a CFX96 Thermal Cycler (BIORAD) based on the protocol described by Scaramozzino-N *et al*. (14). Finally, qPCR data were collected and analyzed using Bio-Rad CFX maestro software (BIORAD). A set of in-house quantified MPXV DNA samples was used as the standard.

### Quantification of infectious titer by Focus Forming Assays (FFA)

 Vero-TMPRSS cells were seeded in a 96-well plate at 4x10^5^ cells/mL. The following day, the monolayer of cells was infected with serially diluted aliquots of cultured viral stocks from each MPXV clade. After 1 hour at 37°C, the media were removed, and a 1:1 ratio of the pre-warmed medium mixed with 2.4% Avicel (DuPont) was overlaid. Plates were incubated at 37°C for 24 hours and then fixed at room temperature for 1 hour using 6% formaldehyde. Cells were permeabilized with 0.1% Triton X-100 for 20 minutes and then blocked with 1% BSA (Sigma-Aldrich) in PBS for 1 hour at room temperature. Subsequently, the samples were incubated with a primary monoclonal antibody targeting the orthopoxvirus A13L protein (Geneva Antibody Facility, AI-582) diluted to 0.25 µg /ml for 1 hour at room temperature. Subsequently, the samples were incubated at room temperature for 30 minutes with a peroxidase-conjugated secondary antibody (Jackson ImmunoResearch) diluted at 1/1500. Foci were visualized by adding True Blue HRP substrate (Avantor). After washing and drying the plates, images were recorded on a Mabtech IRIS Fluorospot/ELIspot reader. A focus was defined as a cluster of adjacent cells expressing the viral antigen. Foci were counted and expressed as FFU/mL.

### Analytical sensitivity testing of cultured virus

Viral stocks from each of the four clades were serially diluted in PBS with a dilution factor of 1:3, beginning at a concentration of 8.00E+08 DNAc/mL. Multiple aliquots of each dilution were prepared for each clade, and the viral load of each dilution was quantified by qPCR (22).

The set of dilutions used to evaluate the POC molecular assays included the lower virus concentrations, whereas the set of dilutions selected for the Ag-RDT evaluation included only the higher concentrations because the expected LOD for Ag-RDTs is much higher than that for POC molecular assays.

All five POC assays were performed according to the manufacturer’s instructions. The only difference was that the virus dilution was added directly to the cartridge (for the two molecular assays) or to the buffer tube (for the three Ag-RDTs) instead of a swab specimen. Each Ag-RDT kit contains its own proprietary buffer. For each dilution prepared for each of the four clades, 200 μL of the dilution was applied in triplicate to the respective buffer of the ACON Biotech and Assure Tech kits while 100 μL of dilution was applied in triplicate to the respective buffer of the SD Biosensor kit. Each manufacturer recommended the input volumes. PBS was used as the negative control. Subsequently, the dilution/buffer mixture was applied to the sample pad of the Ag-RDT under BSL-3 conditions using the materials provided in the kit. The results were independently read by two individuals and interpreted according to the manufacturer’s instructions.

## Data Availability

All data produced in the present work are contained in the manuscript

## Acknowledgement

We thank FIND and the World Health Organization (WHO) for providing the diagnostic tests evaluated in this study. We thank the WHO BioHub, Spiez, for sharing mpox virus isolates. We gratefully acknowledge Francisco Perez Rodriguez, Adriana Renzoni, Valentin Chudzinski and Florian Laubscher of the University Hospitals of Geneva for their contribution to the reference PCR protocol and the sequencing and analysis of the MPXV clade Ib cultured virus. We also extend our sincere thanks to the patients and clinical partners who agreed to share the initial mpox clinical samples, without which this analytical study would not have been possible.

## Supplementary material

**Table S1.** Analytical testing results with clade Ia viral dilutions for the two MPXV POC molecular assays.

| Dilution N° | Reference PCR |  | Xpert® Mpox |  | Standard M10 OPX/MPX |  |
| --- | --- | --- | --- | --- | --- | --- |
|  | DNA copies/mL* | CT | CT MPX | CT OPX | CT MPX | CT OPX |
| 1 | 8.3E+02 | 36.9 | Neg | 37.0 | 30.9 | 31.2 |
|  |  |  | Neg | 37.3 | 30.8 | 31.0 |
|  |  |  | Neg | 37.0 | 30.6 | 31.3 |
| 2 | 2.8E+02 | 38.6 | Neg | 39.4 | 34.1 | 33.1 |
|  |  |  | Neg | 39.7 | 35.0 | 32.9 |
|  |  |  | Neg | 40.2 | 33.0 | 34.9 |
| 3 | 6.5E+01 | 40.4 | Neg | 41 | 34.8 | Neg |
|  |  |  | Neg | 42.4 | Neg | 34.7 |
|  |  |  | Neg | 40.6 | 33.1 | 36.8 |
| 4 | 2.2E+01* | Neg | Neg | 41 | Neg | 36.7 |
|  |  |  | Error |  | Neg | 36.2 |
|  |  |  | Neg | Neg | 35.1 | Neg |
| 5 | 7.3E+00* | Neg | Neg | Neg | Neg | Neg |
|  |  |  | Neg | Neg | Neg | Neg |
|  |  |  | Neg | Neg | 35.3 | Neg |
| 6 | 2.4E+00* | Neg | Neg | Neg | Neg | Neg |
|  |  |  | Neg | Neg | Neg | Neg |
|  |  |  | Neg | Neg | Neg | Neg |

\*When one or more replicates were detected as negative by the reference PCR, the concentration of the viral dilution was estimated based on the dilution factor applied between each step (1:3).

**Table S2.** Analytical testing results with clade Ib viral dilutions for the two MPXV POC molecular assays.

| Dilution N° | Reference PCR |  | Xpert® Mpox |  | Standard M10<br>OPX/MPX |  |
| --- | --- | --- | --- | --- | --- | --- |
|  | DNA copies/mL* | CT | CT MPX | CT OPX | CT MPX | CT OPX |
| 1 | 1.7E+03 | 36.2 | Neg | 35.3 | 29.5 | 31.1 |
|  |  |  | Error |  | 28.2 | 29.1 |
|  |  |  | Neg | 35.3 | 29.8 | 30.9 |
| 2 | 6.8E+02 | 38.3 | Neg | 37.7 | 31.0 | 33.7 |
|  |  |  | Neg | 36.1 | 32.0 | 32.1 |
|  |  |  | Neg | 36.9 | 31.2 | 32.3 |
| 3 | 1.0E+02 | 40.4 | Error |  | 31.9 | 33.2 |
|  |  |  | Neg | 39.3 | 34.8 | 35.7 |
|  |  |  | Neg | 40.4 | 32.1 | 32.5 |
| 4 | 5.1E+01 | 41.6 | Neg | 40.8 | 33.9 | 33.6 |
|  |  |  | Neg | 40.0 | 34.0 | Neg |
|  |  |  | Neg | 39.2 | 35.1 | 36.4 |
| 5 | 1.7E+01* | Neg | Neg | Neg | Neg | Neg |
|  |  |  | Neg | Neg | Neg | Neg |
|  |  |  | Neg | Neg | 35.9 | Neg |
| 6 | 5.7E+00* | Neg | Neg | Neg | Neg | Neg |
|  |  |  | Neg | Neg | Neg | Neg |
|  |  |  | Neg | Neg | Neg | Neg |

*\*When one or more replicates were detected as negative by the reference PCR, the concentration of the viral dilution was estimated based on the dilution factor applied between each step (1:3).*

**Table S3.**
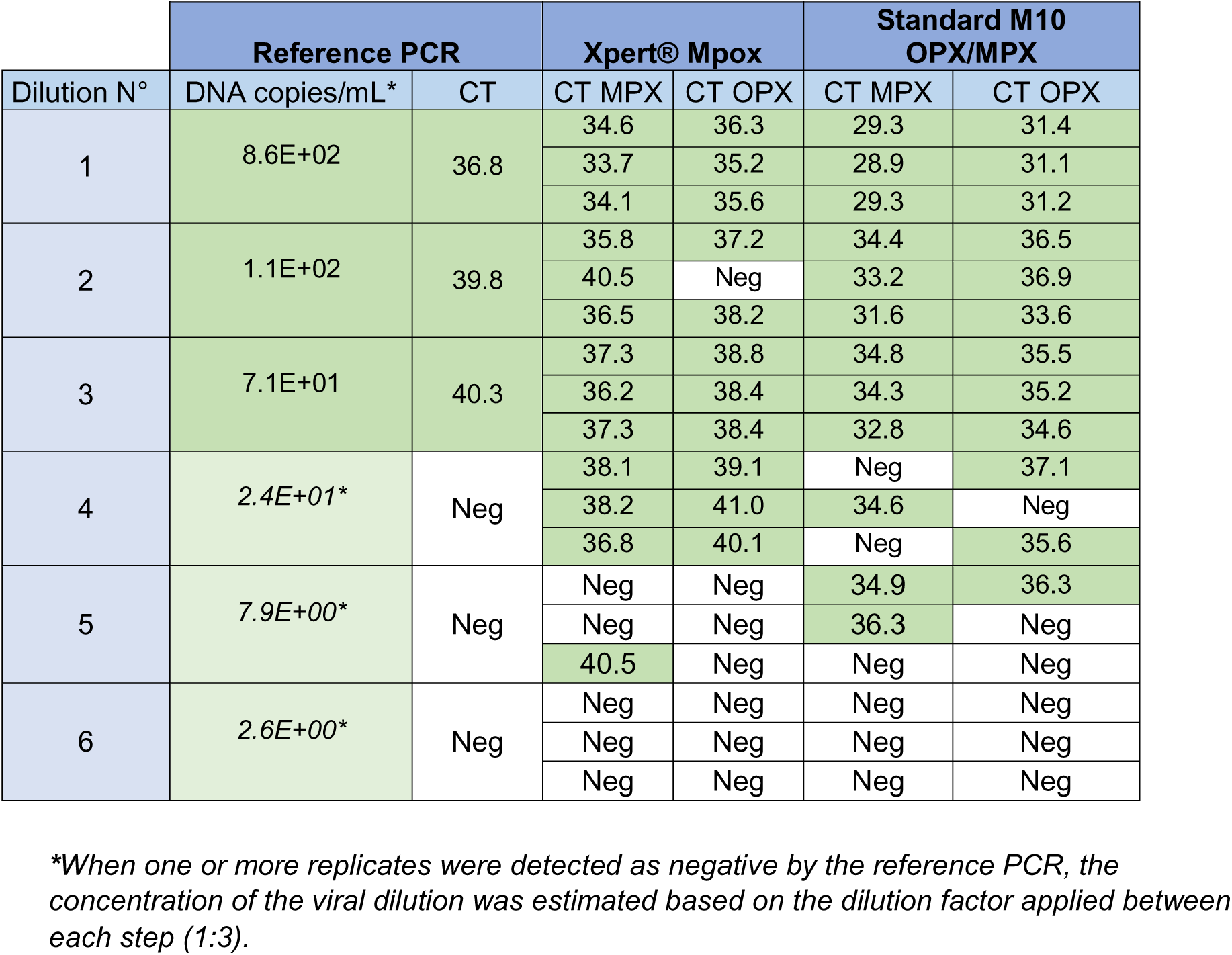
Analytical testing results with clade IIa viral dilutions for the two MPXV POC molecular assays.

| Dilution N° | Reference PCR |  | Xpert® Mpox |  | Standard M10<br>OPX/MPX |  |
| --- | --- | --- | --- | --- | --- | --- |
|  | DNA copies/mL* | CT | CT MPX | CT OPX | CT MPX | CT OPX |
| 1 | 8.6E+02 | 36.8 | 34.6 | 36.3 | 29.3 | 31.4 |
|  |  |  | 33.7 | 35.2 | 28.9 | 31.1 |
|  |  |  | 34.1 | 35.6 | 29.3 | 31.2 |
| 2 | 1.1E+02 | 39.8 | 35.8 | 37.2 | 34.4 | 36.5 |
|  |  |  | 40.5 | Neg | 33.2 | 36.9 |
|  |  |  | 36.5 | 38.2 | 31.6 | 33.6 |
| 3 | 7.1E+01 | 40.3 | 37.3 | 38.8 | 34.8 | 35.5 |
|  |  |  | 36.2 | 38.4 | 34.3 | 35.2 |
|  |  |  | 37.3 | 38.4 | 32.8 | 34.6 |
| 4 | 2.4E+01* | Neg | 38.1 | 39.1 | Neg | 37.1 |
|  |  |  | 38.2 | 41.0 | 34.6 | Neg |
|  |  |  | 36.8 | 40.1 | Neg | 35.6 |
| 5 | 7.9E+00* | Neg | Neg | Neg | 34.9 | 36.3 |
|  |  |  | Neg | Neg | 36.3 | Neg |
|  |  |  | 40.5 | Neg | Neg | Neg |
| 6 | 2.6E+00* | Neg | Neg | Neg | Neg | Neg |
|  |  |  | Neg | Neg | Neg | Neg |
|  |  |  | Neg | Neg | Neg | Neg |

*\*When one or more replicates were detected as negative by the reference PCR, the* *concentration of the viral dilution was estimated based on the dilution factor applied between* *each step (1:3).*

**Table S4.** Analytical testing results with clade IIb viral dilutions for the two MPXV POC molecular assays.

| Dilution N° | Reference PCR |  | Xpert® Mpox |  | Standard M10<br>OPX/MPX |  |
| --- | --- | --- | --- | --- | --- | --- |
|  | DNA copies/mL* | CT | CT MPX | CT OPX | CT MPX | CT OPX |
| 1 | 8.1E+02 | 36.9 | 34.2 | 36.0 | 29.7 | 30.8 |
|  |  |  | 34.4 | 36.0 | 29.9 | 31.7 |
|  |  |  | 34.1 | 36.7 | 29.7 | 31.0 |
| 2 | 3.1E+02 | 38.6 | 35.8 | 37.8 | 30.9 | 32.4 |
|  |  |  | 36.3 | 38.5 | 31.8 | 34.1 |
|  |  |  | 35.3 | 37.5 | 32.5 | 34.5 |
| 3 | 1.7E+02 | 39.0 | 38.1 | 38.1 | 32.4 | 35.5 |
|  |  |  | 38.2 | 40.0 | Neg | 35.6 |
|  |  |  | 38.0 | 38.7 | 33.1 | 35.1 |
| 4 | 1.0E+02 | 39.1 | Neg | 41.3 | Neg | Neg |
|  |  |  | 39.3 | 40.7 | 33.4 | 33.5 |
|  |  |  | Error |  | 34.1 | 35.5 |
| 5 | 3.4E+01* | Neg | Neg | Neg | 35.1 | Neg |
|  |  |  | Neg | Neg | Neg | 35.6 |
|  |  |  | 40.2 | Neg | 36.1 | 36.6 |
| 6 | 1.1E+01* | Neg | Neg | Neg | Neg | Neg |
|  |  |  | 40.2 | Neg | Neg | Neg |
|  |  |  | 40.5 | 42.4 | Neg | Neg |

*\*When one or more replicates were detected as negative by the reference PCR, the concentration of the viral dilution was estimated based on the dilution factor applied between each step (1:3).*

**Table S5.**
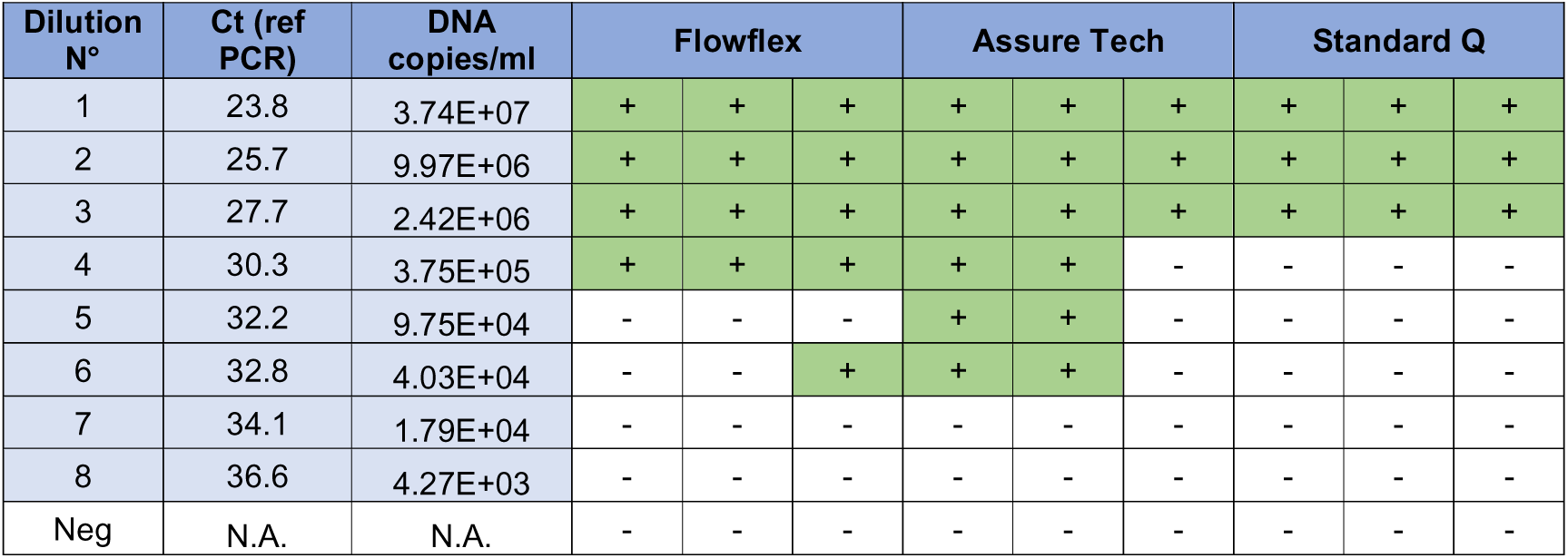
Analytical testing results with clade Ia viral dilutions for the three MPXV antigen rapid tests.

**Table S6.**
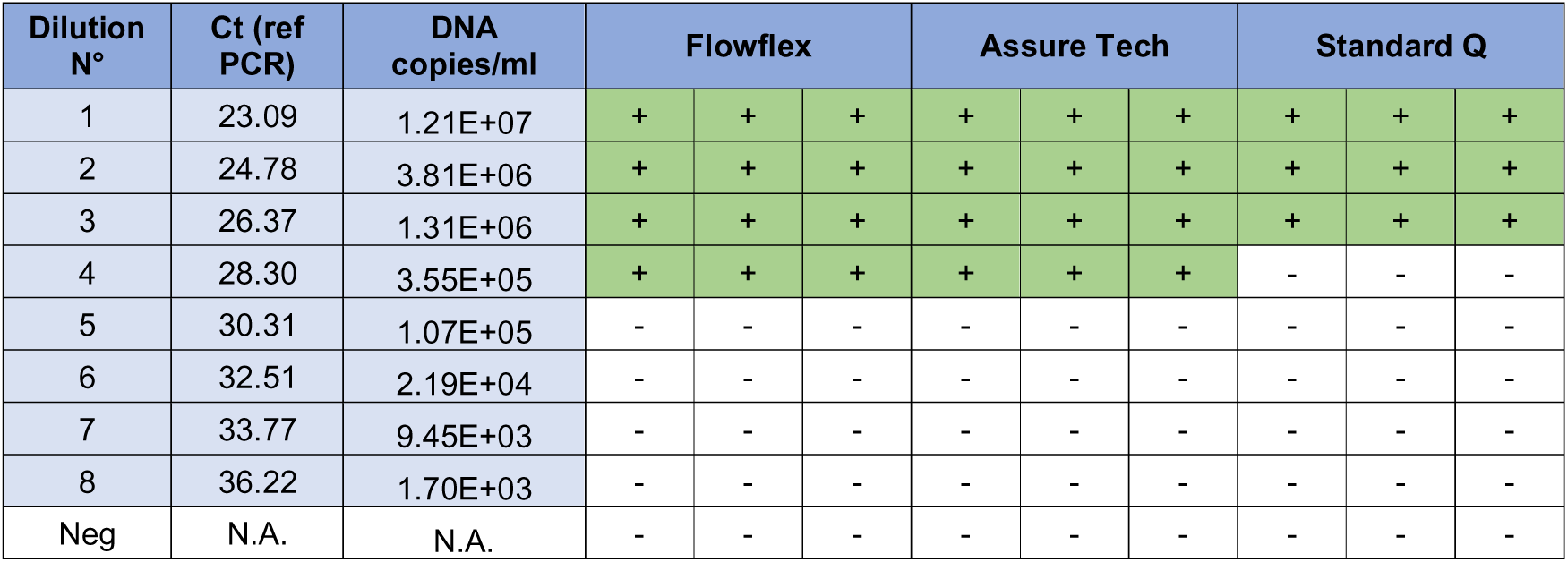
Analytical testing results with clade Ib viral dilutions for the three MPXV antigen rapid tests.

**Table S7.**
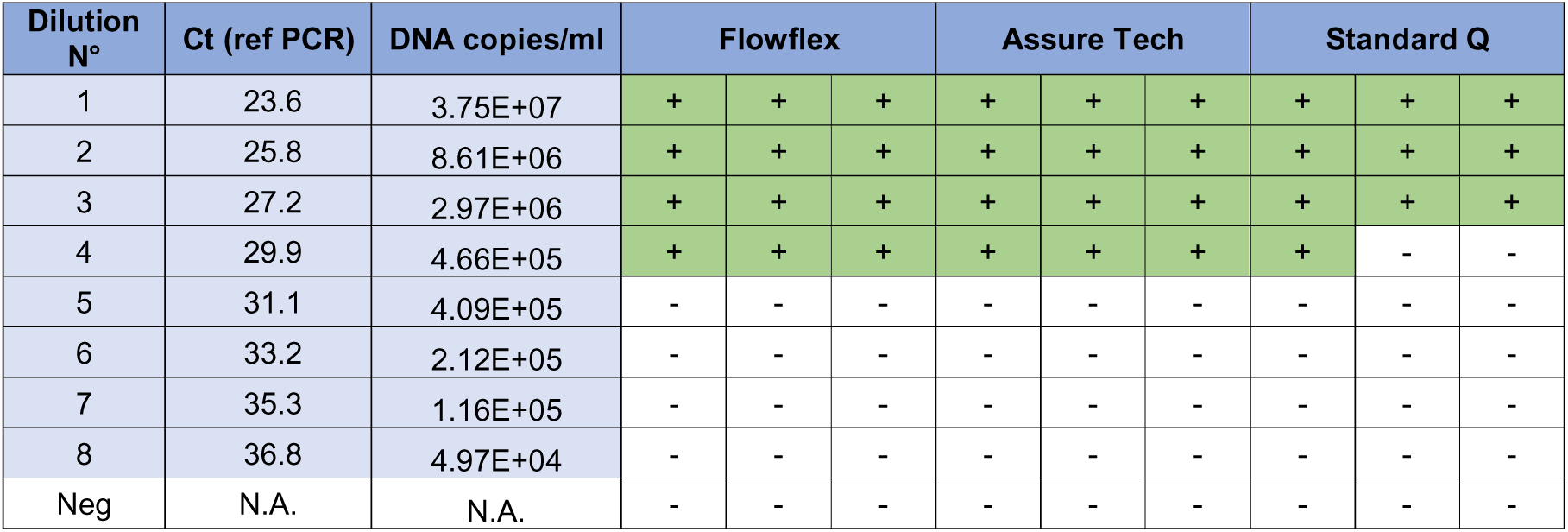
Analytical testing results with clade IIa viral dilutions for the three MPXV antigen rapid tests.

**Table S8.** Analytical testing results with clade IIb viral dilutions for the three MPXV antigen rapid tests.

| Dilution N° | Ct (ref PCR) | DNA copies/ml | Flowflex |  |  | Assure Tech |  |  | Standard Q |  |  |
| --- | --- | --- | --- | --- | --- | --- | --- | --- | --- | --- | --- |
| 1 | 24.5 | 2.08E+07 | + | + | + | + | + | + | + | + | + |
| 2 | 25.9 | 7.93E+06 | + | + | + | + | + | + | + | + | + |
| 3 | 29.1 | 8.37E+05 | + | + | + | + | + | - | + | + | + |
| 4 | 30.1 | 4.18E+05 | - | - | - | + | - | - | - | - | - |
| 5 | 31.5 | 1.62E+05 | - | - | - | - | - | - | - | - | - |
| 6 | 34.2 | 2.52E+04 | - | - | - | - | - | - | - | - | - |
| 7 | 36.9 | 4.35E+03 | - | - | - | - | - | - | - | - | - |
| 8 | 39.4 | 8.09E+02 | - | - | - | - | - | - | - | - | - |
| Neg | N.A. | N.A. | - | - | - | - | - | - | - | - | - |

**Table S9.** References of the DNA sequences of the four MPXV strains used for the viral dilutions and analytical sensitivity testing.

| Sequence Definition | Organism | MPXV Clade | GenBank accession number |
| --- | --- | --- | --- |
| Monkeypox virus isolate CMVE-1, partial genome | Monkey pox virus | Clade Ia | PX049154 |
| Monkeypox virus isolate CMVE-2, partial genome | Monkey pox virus | Clade Ib | PX049155 |
| Monkeypox virus isolate CMVE-3, partial genome | Monkey pox virus | Clade IIa | PX049156 |
| Monkeypox virus isolate hMPXV-CH-38347314/2022, partial genome | Monkey pox virus | Clade IIb | OP783902 |

**Figure S1.**
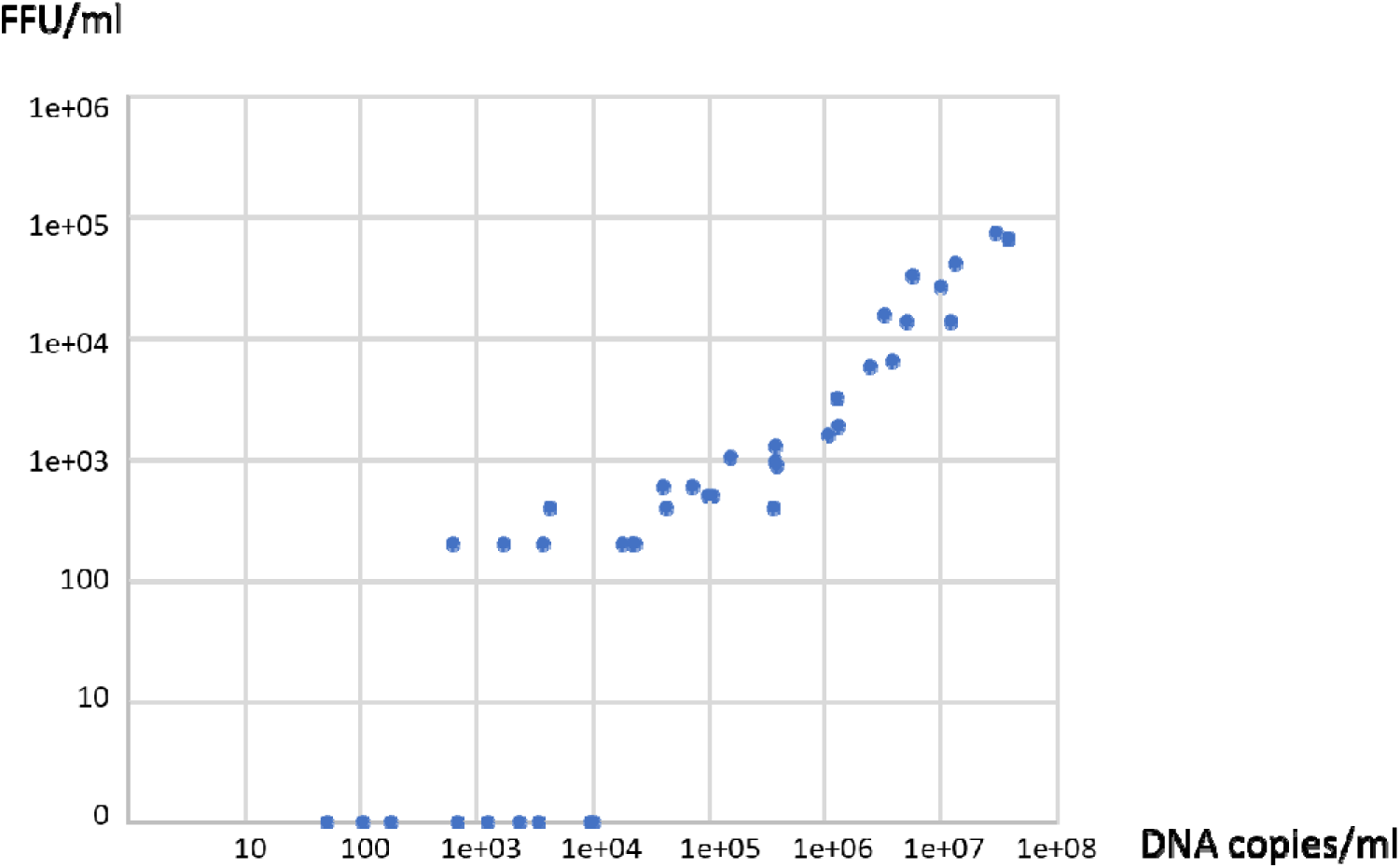
Scatter plot of measured DNA concentrations in DNA copies/mL vs FFU/mL for all dilutions titrated by Focus Forming Assay (FFA)

